# Mechanistic rationale of drugs, Primary endpoints, Geographical distribution of clinical trials against Severe acute respiratory syndrome-related coronavirus-2: A Systematic review

**DOI:** 10.1101/2020.05.24.20112169

**Authors:** Bhanu Prasad Venkatesulu, Thoguluva Chandrasekar Viveksandeep, Prashanth Girdhar, Pragathee, K Patel, K Harsh, Jacob Manteuffel

## Abstract

**Objective:** To do a systematic review and critical appraisal of the ongoing clinical trials that are assessing various therapeutic interventions against SARS-CoV-2 with an aim to provide insight into the various interventions tested, clinical rationale, geographical distribution of the trials as well as the endpoints assessed in the studies.

**Design:** Rapid systematic review and critical appraisal of the ongoing clinical trials against SARS-CoV-2.

**Data sources:** ClinicalTrials.gov, World health organization (WHO) International Clinical Trials Registry Platform (ICTRP) and Cochrane COVID registry were assessed till May 11^th^ 2020.

**Study selection:** Studies on any intervention based randomized controlled trials (RCTs), prospective clinical studies on SARS-CoV-2 in patients ≥18 years of age. Studies on autopsy series, preclinical studies, diagnostic methods, mathematical modelling, epidemiology and health services research, pediatric populations were excluded.

**Data extraction:** The data was extracted by two authors independently into pre-defined forms based on the SPIRIT 2013 checklist. The data was extracted on various domains such as trial number, study title, abstract of the study, interventions assessed, sample size, phase of the study, study sponsor, primary endpoint assessed and country of study.

**Results:** The search resulted in 3242 ongoing studies of which 829 studies were included. There are 134 different drug-based interventions being assessed in 463 clinical trials as treatment options. Seventy-two studies assessed preventive options of which 53 are drug-based prophylaxis and 19 assessed vaccines. Herbal medicines are being assessed in 79 studies; convalescent plasma therapy in 56 studies; stem cell-based interventions in 42 studies; anesthesia-based interventions in 31 studies, machine-based interventions in 24 studies, mental health-based interventions in18 studies, rehabilitation-based interventions in 12 studies and miscellaneous interventions in 32 studies. China accounts for 35% of all ongoing clinical studies followed by USA 23%, France 7%, Spain 3.3%,Canada 2%, multi-country studies account only for 1.5% (13) and other countries together account for 28%.Amongst the 463 studies assessing drug-based treatment options, studies that are funded by federal and academic institutions are 79.6%, pharmaceutical company funded studies are 15.11% and no funding information is available in 5.10%. The definitive outcomes like mortality are being assessed as primary outcome in 22.8% of the studies only and need for ventilator in 6.2% of the studies. Rest of the studies has primary outcomes such as clinical recovery (15.9%), viral clearance(17.4%), time to recovery (10.1%), oxygen improvement (5.6%), ICU admission (1.9%), lab and imaging(6.4%), adverse effects (5.3%) and symptom reduction(1.5%),no outcome reported(6.2%) which account for 71% of the studies. Amongst the pharmaceutical company funded drug-based studies, only 20% of the studies had mortality as the primary outcome. Only 5.5% of the ongoing clinical trials are specifically designed to assess the most vulnerable population like elderly, patients with comorbidities and cancer. The most common intervention being tested against COVID-19 are antimalarial medications with 105 clinical studies. Hydroxychloroquine is the most common drug being tested with 83 ongoing studies.

**Conclusion:** Multiple intervention based clinical studies against SARS-CoV-2 are being performed throughout the world with a high concentration of clinical trials in the developed world. There is a high concern that most of the studies maybe repetitive; elderly and patients with comorbidities are being underrepresented; definite endpoints like mortality are being assessed in only one-fifth of the studies.

## Introduction

Severe acute respiratory syndrome-related coronavirus 2 (SARS-CoV-2) is the causative agent for coronavirus disease (COVID-19) that has led to nearly 5 million new cases and more than a quarter million deaths as of May 22, 2020 (1). The case fatality rate is significantly higher in elderly, patients with preexisting comorbidities like hypertension, diabetes, cardiovascular diseases chronic respiratory disease and for cancer(2).The high rate of transmission, moderate case fatality, novel nature of the virus has made the virus a formidable pathogen and has left a huge burden on the health care infrastructure.

Enormous amount of research is ongoing for finding vaccines, therapeutic interventions to prevent, mitigate, treat and manage the complications of COVID-19 disease. Randomized clinical trials (RCT) form the backbone of evidence-based rationale approach for management of any disease. SARS-CoV-2 pandemic has led to a flurry of clinical trials being performed throughout the world. The interventions being tested are largely based on known antiviral activity against SARS and MERS (Middle Eastern Respiratory Syndrome), efficacy in the invitro and in vivo models of SARS-CoV-2, potential docking sites on the viral genome based on computational modelling studies and biological agents to counter the cytokine surge and withspread immune activation(3, 4).There has also been a surge in interest in repurposing previously approved FDA drugs like Ivermectin, Chlorpromazine, Isotretinoin, Nitazoxanide for possible anti-viral activity(5, 6). The simultaneous initiation of multiple clinical trials has led to redundancy of study design, lack of novelty and absence of pragmatic primary end points. We undertook this systematic review of ongoing interventional clinical trials to collate the available information on the study design, geographical distribution, endpoints assessed and various drugs that are being assessed in the fight against SARS-CoV-2 infections.

## Methods

This systematic review has been performed and reported in compliance with the Preferred Reporting Items for Systematic Reviews and Meta-Analyses(7).(**Supplementary data**)

### Search strategy

ClinicalTrials.gov, World health organization (WHO) International Clinical Trials Registry Platform (ICTRP) and Cochrane COVID registries were assessed up to May 11th with the search terms coronavirus, SARS-CoV-2 by two independent investigators (BV, VT). The clinical trials from the initial search of the electronic databases were imported into reference manager software. An independent review of the clinical trials was done. The duplicates were removed and the titles of the clinical trials were evaluated. Trials relevant to the topic of interest were shortlisted. The clinical studies that fulfilled the inclusion criteria were shortlisted for final systematic review. Reasons for excluding clinical studies were documented.

### Inclusion and exclusion criteria

#### Study selection

The following eligibility criteria were used:

*Inclusion criteria:* (i) Any intervention based randomized controlled trials (RCTs), prospective clinical study on SARS-CoV-2 ii) Patients ≥18 years of age.

*Exclusion criteria:* (i) Autopsy series, preclinical studies ii) studies reporting diagnostic methods, mathematical modelling, epidemiology and health services research iii) Studies in pediatric populations iv) Studies on SARS-CoV and MERS

### Data extraction

The data was extracted by two authors independently using a standardized data extraction form based on the SPRINT checklist. The data was extracted on various domains such as trial number, study title, abstract of the study, interventions assessed, sample size, phase of the study, study sponsor, primary endpoint assessed and country where study is being done. Any discrepancies in extraction of data was resolved by mutual discussion (BV, VT). Details on data extraction are provided in the **Supplementary data**.

### Definitions

We stratified studies into intervention-based studies, observational studies, mathematical modelling studies, studies assessing various diagnostic methods for SARS-CoV-2, studies that assess health services research, epidemiological studies, studies on pediatric population.

## Results

### Study search and study characteristics

The search of the clinical trial databases resulted in 3242 ongoing studies of which 913 underwent full review and 829 studies were included in the systematic review (**figure 1**). Amongst the 829 articles, 463 assessed various drugs as treatment options against COVID-19; 72 studies assessed preventive options of which 53 are drug-based prophylaxis and 19 studies assessed vaccines; herbal medicines are being assessed in 79 studies; convalescent plasma therapy is being studied in 56 studies; stem cell-based interventions in 42 studies; anesthesia-based interventions in 31 studies, machine-based interventions in 24 studies, mental health-based interventions in18 studies; rehabilitation-based interventions in 12 studies; miscellaneous interventions in 32 studies. China accounts for 35% (291) of all ongoing clinical studies followed by USA 23% (188), France 7% (63), Spain 3.3% (28), Canada 2% (20), multi-country studies account only for 1.5% (13) and other countries together account for 28% of the studies (**Supplementary data)**. There are only 2.77% of clinical trials that specifically seek to enroll patients with comorbidities like Diabetes, hypertension, cardiac disease; 2% of the trials that are being done in the elderly and 0.70% in cancer patients accounting for a total of 5.5% ongoing clinical trials.

**Figure 1:**
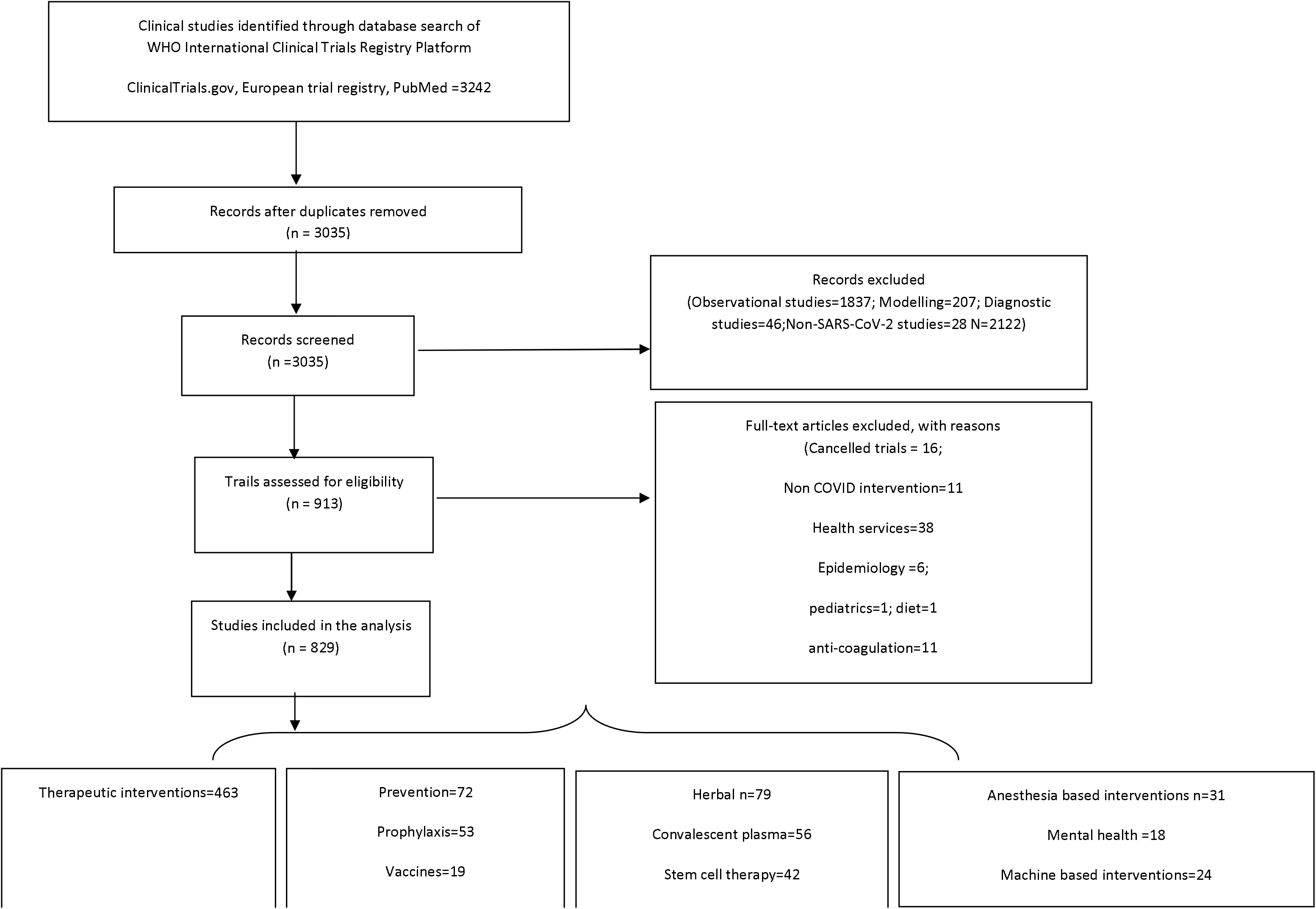
PRISMA flow diagram depicting the search strategy utilized in the systematic review

### Drug based interventions as treatment against COVID-disease

There are 134 different drug-based interventions being assessed in 463 clinical trials throughout the world (**Table 1)**. Amongst the 463 studies assessing drug-based treatment options, studies that are funded by federal and academic institutions are 79.6%; pharmaceutical company funded studies are 15.11% (70); no funding information has been provided in 5.10% (24) studies. The definitive outcomes like mortality was assessed as primary outcome in 22.8% of the studies only and need for mechanical ventilation in 6.2% of the studies. Rest of the studies had outcomes such as: clinical recovery (15.9%), viral clearance (17.4%), time to recovery (10.1%), oxygen improvement (5.6%), ICU admission (1.9%), labs and imaging(6.4%), adverse effects (5.3%) and symptom reduction(1.5%),no outcome reported(6.2%) which account for 71% of the studies. Amongst the pharmaceutical company funded drug-based studies, only 20% of the studies had mortality as the primary outcome and 7% had need for mechanical ventilation as an outcome.

**Table 1:**
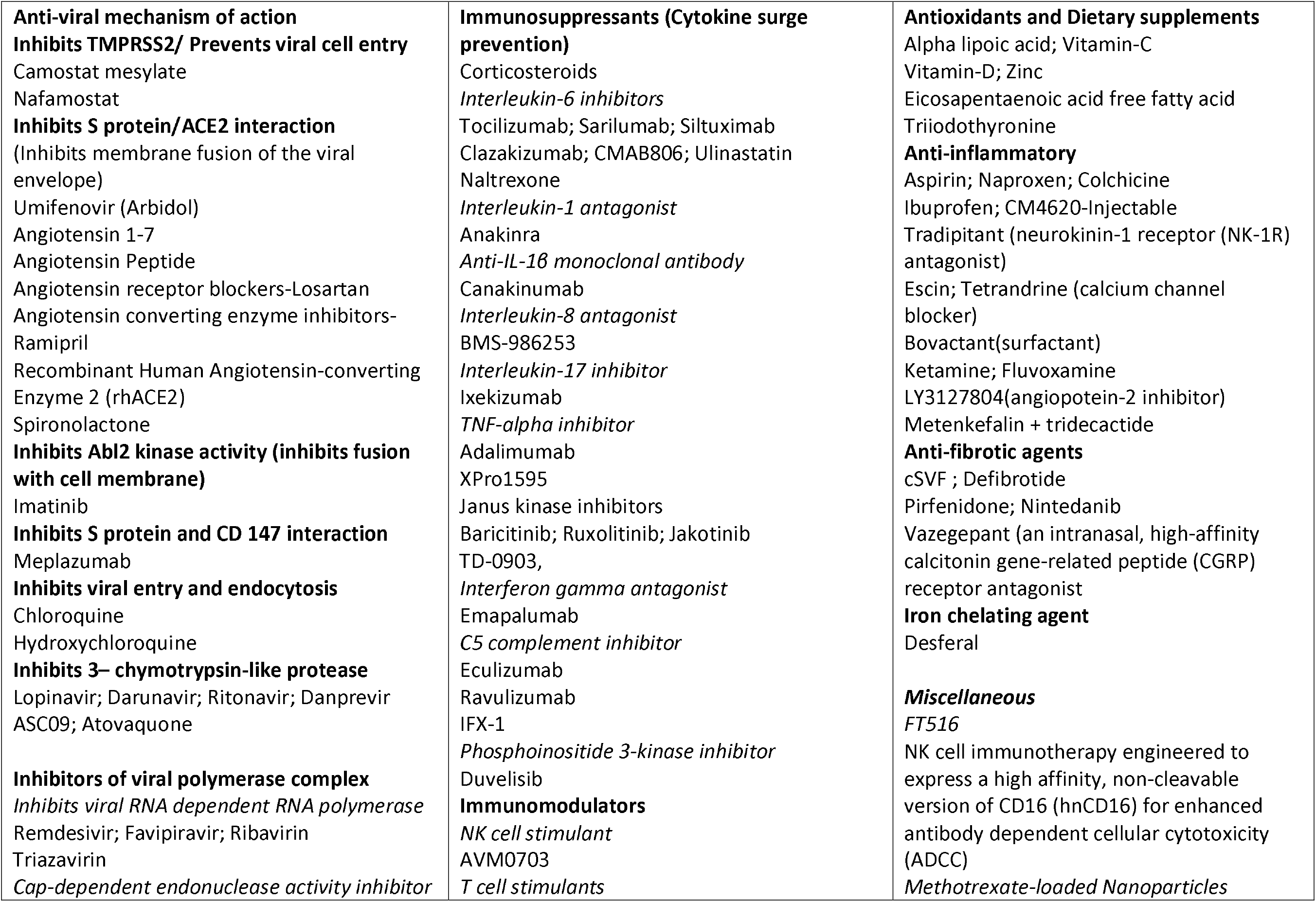
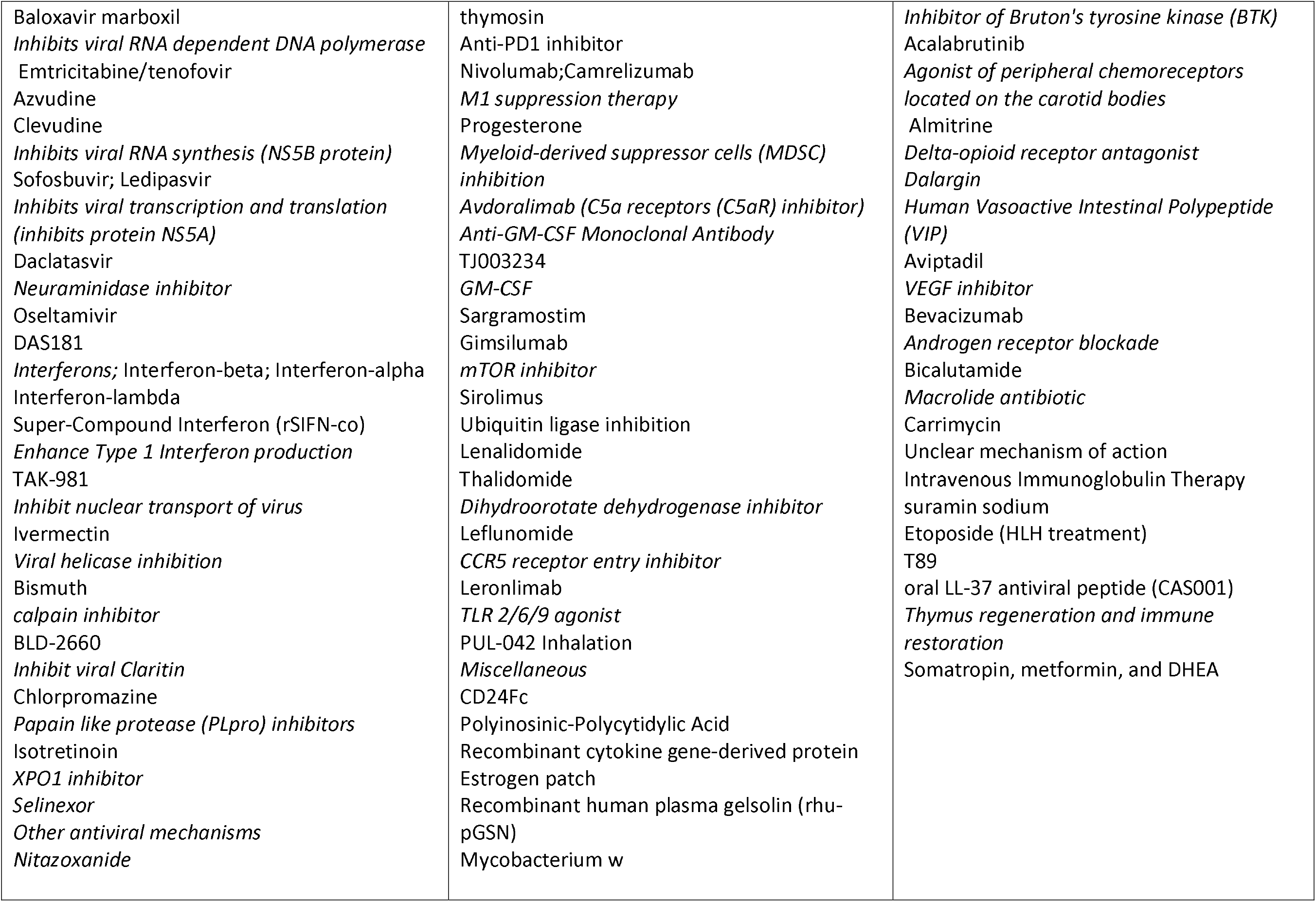
Summary of mechanisms of the drugs used in the clinical trials as treatment against SARS-CoV-2 infection.

The most common drug-based treatment intervention being tested against COVID-19 are antimalarial medications with 105 clinical studies. Hydroxychloroquine is the most common drug being tested with 83 ongoing studies of which hydroxychloroquine alone is being studied in 49 studies and hydroxychloroquine with azithromycin being studied in 22; Chloroquine is being tested in 31 studies. Antiviral medications are being tested in 76 clinical trials of which Lopinavir with Ritonavir in 28 studies, Favipravir in 13 studies, Remdesivir in 8 studies, interferons in 14 studies; other antivirals in 13 studies. Immunosuppressants are being assessed in 82 studies-Interleukin-6 antagonists in 38 studies of which Tocilizumab based are 23 studies, Sarilumab in 7 studies; Corticosteroids in 15 studies; Immunomodulators in 30 studies; Anti-inflammatory agents in 20 studies colchicine in 8 studies; Anti-oxidants and dietary supplements like vitamin C, D, Zinc in 16 studies; Antifibrotic agents in 9 studies; other miscellaneous interventions account for the rest of the studies. These studies are most commonly done in China (122) followed by USA (114) and France (47) (**Supplementary data)**. The largest ongoing clinical trial is the WHO sponsored Solidarity trail which is a multi-center study with a sample size of 1,00,000 patients to assess Remdesivir, chloroquine or hydroxychloroquine, lopinavir plus ritonavir, interferon-beta with control, with a primary endpoint of all-cause mortality(8, 9). NCT04292899 is a multi-center study with a sample size of 6,000 that is assessing Remdesivir with a primary endpoint of improvement on a 7-point Ordinal Scale on Day 14 (10). NCT04322682 is a Canadian study that is assessing Colchicine with a primary endpoint of all-cause mortality at day 30(11). **Table 2** provides a summary of phase 3 clinical trials with sample size of more than thousand patients that assess various interventions in COVID-19 disease.

**Table 2:** Summary of drug-based treatments in phase 3 clinical trials with sample size of more than thousand patients against SARS-CoV-2.

| Study identification number | Country | Sample size | Intervention assessed | Primary endpoint |
| --- | --- | --- | --- | --- |
| NCT04292899 | Multicenter study; sponsor-Gilead | 6000 | Remdesivir | The Odds of Ratio for Improvement on a 7-point Ordinal Scale on Day 14 |
| NCT04292730 | Multicenter study; sponsor-Gilead | 1600 | Remdesivir | The Odds of Ratio for Improvement on a 7-point Ordinal Scale on Day 11 |
| NCT04315948 | Multicenter study; sponsor-National institute of France | 3000 | Remdesivir; Lopinavir/ritonavir; Interferon Beta-1A; Hydroxychloroquine | Percentage of subjects reporting each severity rating on a 7-point ordinal scale on day 15 |
| NCT04324047 | France<br>Sponsor- Assistance Publique - Hôpitaux de Paris | 1000 | immune modulator drugs | Overall survival at day 14<br>WHO progression scale COVID 19 |
| NCT04322682 | Canada<br>Sponsor-Montreal heart institute | 6000 | Colchicine | Number of participants who die or require hospitalization due to COVID-19 infection upto day 30 |
| ISRCTN83971151<br>Solidarity trial | Multicenter study<br>Sponsor-WHO | 100000 | Remdesivir, chloroquine or hydroxychloroquine, lopinavir plus ritonavir, and interferon-beta. | All-cause mortality, subdivided by the severity of disease at the time of randomization, measured using patient records throughout the study |
| NCT04328012 | USA<br>Sponsor- Bassett Healthcare | 4000 | lopinavir/ritonavir; Hydroxychloroquine Sulfate; Losartan | National Institute of Allergy and Infectious Diseases COVID-19 Ordinal Severity Scale (NCOSS) at 60 days |
| NCT04345289 | Denmark<br>Sponsor-Thomas Benfield | 1500 | Convalescent anti-SARS-CoV-2 plasma; Sarilumab; Baricitinib; Hydroxychloroquine | All-cause mortality or need of invasive mechanical ventilation at day 28 |
| NCT04358068 | USA<br>Sponsor-NIH | 2000 | Hydroxychloroquine +azithromycin | Proportion of participants who died from any cause or were hospitalized at day 21 |
| NCT04356495 | France<br>Sponsor- University Hospital, Bordeaux | 1057 | Hydroxychloroquine; Imatinib; Favipiravir; Telmisartan | Proportion of participants with an occurrence of hospitalization or death at day 14 |
| NCT04358003 | USA sponsor- Marker Therapeutics AG | 2000 | Plasma Adsorption Cartridge | All-cause mortality at day 28 |

### Drug being tested as prophylaxis against COVID-disease

Fifty-three drug-based studies are being assessed as prophylaxis in COVID-19.HCQ is the most commonly assessed prophylaxis drug being studied in 24 clinical studies; chloroquine in 2 studies; HCQ+ Azithromycin in 1 study; Chloroquine+ Azithromycin in 1 study; Nitazoxamide in 2 studies; other anti-viral medications include Camostat mesylate, interferons, Lopinavir + Ritonavir. Other immunomodulatory medications being assessed include anakinra, colchicine, corticosteroids, Levamisole, mycobacterium W in one study each. **Table 3** provides a summary of the prophylactic interventions being assessed against COVID-19. The most common country where these studies are conducted are USA (17) followed by 4 each in France and China (**Supplementary data)**. The largest of the prophylactic study is the Crown coronation study which is a multicenter study with a sample size of 55,000 that is assessing various doses of chloroquine in the effectiveness in preventing laboratory-confirmed symptomatic COVID-19 in healthcare workers with repeated exposures to SARS-CoV-2(12).The next largest study is the CRASH 19 study conducted in UK with an sample size of 10,000 that is assessing Aspirin, Losartan and Simvastatin with the all-cause mortality as the primary endpoint(13). WHIPCOVID study is a multicenter study assessing efficacy of weekly vs daily HCQ in preventing new cases of SARS-CoV-2 infection(14).

**Table 3:** Summary of preventive phase 3 clinical trials with sample size of more than thousand patients against SARS-CoV-2 infection.

| Study identification number | Country | Sample size | Intervention assessed | Primary endpoint |
| --- | --- | --- | --- | --- |
| NCT04333407<br>Phase 3 trial | United Kingdom<br>Sponsor-Imperial College London | 3170 | Aspirin; Clopidogrel; Rivaroxaban; Atorvastatin; Omeprazole | All-cause mortality at 30 days after admission |
| NCT04333732<br>Phase 3 trial<br>Crown coronation study | Multicenter study<br>Sponsor- Bill and Melinda Gates Foundation | 55000 | Different doses of chloroquine | Effectiveness in preventing laboratory-confirmed symptomatic COVID-19 in healthcare workers with repeated exposures to SARS-CoV-2. |
| NCT04334967<br>Phase 3 trial | USA<br>Sponsor- Providence Health & Services | 1250 | Hydroxychloroquine | Percentages of enrolled patients needing hospitalization and mechanical ventilation at day 14 |
| NCT04328467<br>Phase 3 trial | USA<br>Sponsor- University of Minnesota | 3500 | Hydroxychloroquine | Outcome reported as the percent of participants in each arm who are COVID-19-free at the end of study treatment upto 12 weeks |
| NCT04341441<br>Phase 3 trial<br>WHIP COVID study | USA<br>Sponsor- Henry Ford Health System | 3500 | Hydroxychloroquine daily vs weekly dosing | Measure the difference in new cases of COVID-19 disease between randomized treatment arms at 8 weeks |
| NCT04342156<br>Phase 3 trial | Singapore<br>Sponsor- Tan Tock Seng Hospital | 1200 | Hydroxychloroquine | Positive serology or reverse transcriptase (RT-PCR) for COVID-19 up until day 28 |
| NCT04343001<br>Phase 3 trial<br>CRASH 19 study | United Kingdom<br>Sponsor-London School of Hygiene and Tropical Medicine | 10000 | Aspirin, Losartan and Simvastatin | Death upto 28 days from day of randomization |

### Vaccines

The most important option to prevent further waves of the COVID pandemic are vaccines. There are 19 vaccine-based studies of which are 8 are BCG and 1 measles vaccine-based studies; 11 are newer vaccines candidates. The most commonly studied vaccine is the BCG vaccine in 8 studies followed by recombinant novel Coronavirus Vaccine (Adenovirus Type Vector) in two studies, aAPC Vaccine, Minigene Vaccine, recombinant chimeric COVID-19 epitope DC vaccine, bacTRL-Spike Vaccine, Measles vaccine, mRNA-1273 vaccine, nanoparticle Vaccine, ChAdOx1 nCoV-19 and INO-4800.China is leading the initiative with four ongoing human trials followed by USA with 3 trials. **Supplementary data** provides a summary of the ongoing vaccine-based studies around the world.

### Mesenchymal stem cell therapy

There are currently 42 clinical studies that are assessing mesenchymal stem cell therapy-based interventions in COVID-19 disease. China is leading the initiative with 25 ongoing human trials followed by USA with 8 trials and Spain with 4 trials. The rationale for use of mesenchymal stem cells are the hypothesized immunomodulatory properties that can counter the cytokine storm. The various sources of stem cells that are being studied include cord blood, human menstrual blood-derived, mesenchymal stem cells exosomes atomization, human dental pulp, human stromal cells, umbilical cord blood mononuclear cells and umbilical cord Wharton’s Jelly derived mesenchymal stem cells (**table 4**).

**Table 4:** Summary of interventions used in the clinical trials as prevention against SARS-CoV-2 infection.

|  |  |
| --- | --- |
| **Prevention**<br>**Pre and post exposure Prophylaxis**<br>*Antiviral medications*<br>Hydroxychloroquine<br>Chloroquine<br>Lopinavir/Ritonavir<br>Interferon alpha<br>Camostat Mesylate<br>Peginterferon Lambda-1a<br>Nitazoxanide<br>*Anti-inflammatory*<br>Anakinra<br>Colchicine<br>Corticosteroids<br>*Immunomodulators*<br>Levamisole and Isoprinosine<br>Lactobacillus Coryniformis K8<br>Lenzilumab<br>*Miscellaneous*<br>Vitamin C; Vitamin D; Zinc<br>Melatonin<br>Mycobacterium w<br>rhIFNa Nasal Drops<br>Resistant Potato Starch<br>Nitric oxide<br>Bêta-cyclodextrin and citrox mouthwash<br>Povidone-iodine 0.5% nasal | **Vaccines**<br>Recombinant Novel Coronavirus Vaccine (Adenovirus Type 5 Vector)<br>aAPC Vaccine<br>Minigene Vaccine<br>Recombinant chimeric COVID-19 epitope DC<br>BCG V<br>bacTRL-Spike<br>Measles<br>RNA Vaccine Candidate<br>mRNA-1273 vaccine<br>Nanoparticle Vaccine<br>Recombinant novel coronavirus (adenovirus type 2 vector)<br>ChAdOx1 nCoV-19<br>INO-4800<br>*Non pharmacological interventions*<br>App based social distancing<br>Face masks vs N95 respirator<br>Internet based solutions<br>Isolation strategy |

**Table 5:**
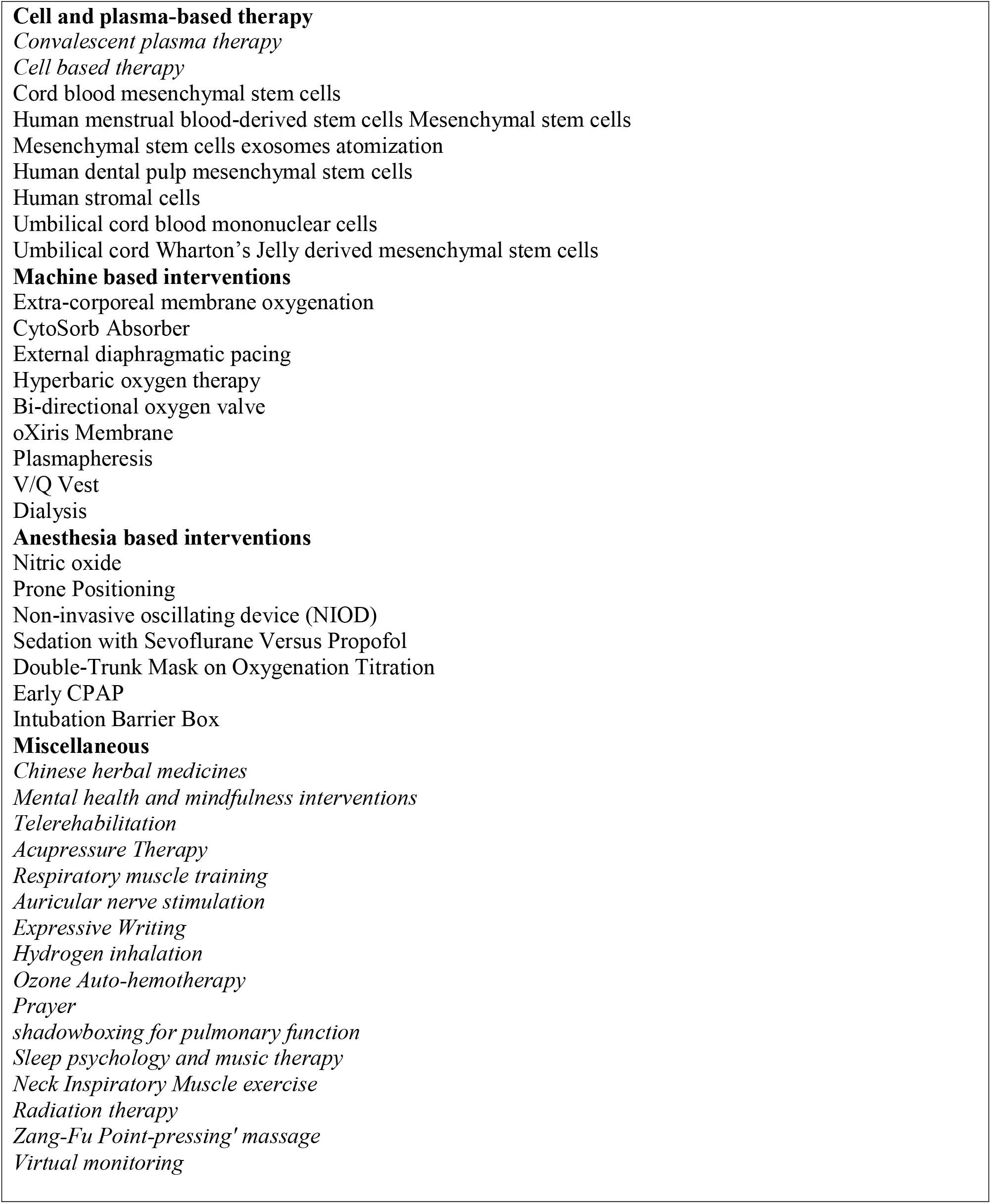
Summary of non-pharmacological interventions used in the clinical trials as treatment against SARS-CoV-2 infection.

### Convalescent plasma therapy

Convalescent plasma therapy (CPT) involves infusion of plasma obtained from people who have recovered from COVID-19 and who have circulating neutralizing antibodies which provides short-term immunity against SARS-CoV-2 coronavirus. There are currently 56 ongoing studies that are assessing CPT with 20 studies being done in USA and 14 studies being done in China. The largest ongoing clinical trial is the CONCOR-1 study being done in Canada with a sample size of 1000 patients with the aim to assess if CPT reduces in-hospital mortality in patients hospitalized for COVID-19. (**table 4**)

### Herbal medicines

There are currently 79 clinical studies assessing the efficacy of alternative medicines mainly the Chinese herbals that are being assessed in 77 studies followed by 1 study in Iran and one Ayurveda study in UK.

### Supplementary data

provides a summary of the various ongoing studies that are testing herbal medicines against COVID-19.

### Anesthesia based interventions

There are currently 31 ongoing studies that assess various anesthesia-based interventions that include prone positioning (11), nitric oxide inhalation (7), ventilator settings (8) and modified intubation techniques (5). USA is performing 7 studies followed by Canada with 5 studies.

### Machine based interventions

There are currently 24 ongoing studies that are assessing machine-based interventions like extra-corporeal membrane oxygenation (ECMO) (8), plasmapheresis(5), CytoSorb Adsorber (3), Bidirectional Oxygenation Valve(2), hyperbaric oxygen therapy(2),continuous renal replacement therapy(2), external diaphragmatic pacing(1), oXiris Membrane (1), and V/Q Vest(1).

### Mental health

There is excess emotional distress to patients and healthcare providers during the COVID-19 pandemic. There are 18 studies that are assessing mental health-based interventions that include mindfulness (11) meditation (3), intelligent psychosomatic adjustment system (2), cognitive behavioral therapy (1), and psychological support. China is performing 8 studies followed by USA with 4 studies.

### Rehabilitation

There are currently 12 studies that are assessing various rehabilitation initiatives like telerehabilitation-based exercises (6), general rehabilitation (2), pulmonary rehabilitation (2) anosmia rehabilitation (1), and rehab -meals (1). China and Turkey are leading with 3 studies each.

### Others

There are 32 other intervention-based studies that were not categorized into the above distinctions. They include microbiota transplantation (3), Natural Killer Cell (CYNK-001) infusions (3), acupressure therapy at auricular point (3), hydrogen inhalation (3), radiation therapy (2), respiratory muscle training (2), digital stress through artificial intelligence(2), Ozone autohemotherapy(2), virtual monitoring(2), expressive writing(1), face mask(1), internet based Solution(1), Medical Masks vs N95 Respirators(1),online distance learning(1), prayer(1),shadowboxing for pulmonary function(1),sleep psychology and music therapy (1),Zang-Fu Point-pressing’ massage(1) and endo-venous systemic Ozone therapy(1).

## Discussion

In this systematic review, we critically appraise 829 ongoing clinical trials that are assessing various interventions like treatment, drug-based prophylaxis, herbal medicines, CPT, stem cell-based interventions, anesthesia-based interventions, machine-based interventions, mental health-based interventions, rehabilitation-based interventions and miscellaneous interventions. China and USA account for majority of the ongoing studies with concern that patients in middle- and low-income countries may have minimal access for enrollment into clinical trials. Multi-center multi-country collaborative studies account only for 1.5% of all ongoing clinical studies which show apparent lack of collaborative effort among researchers as well as difficult in universal applicability of the conclusions made from ongoing studies done in developed countries.

The major overwhelming nature of this pandemic has been case fatality rates of 1-10% seen in different healthcare settings and the high demand for ventilatory support. Despite this,71% of the clinical trials have surrogate endpoints other than all-cause mortality or ventilatory support as the primary endpoint. In addition, the case fatality rate is significantly higher in elderly, patients with preexisting comorbidities like hypertension, diabetes, cardiovascular diseases, chronic respiratory disease and cancer. Despite this, only 5.5% of the ongoing clinical trials specifically seeks to enroll this subgroup of patients. This causes significant concern regarding applicability of the ongoing clinical trials to the general population and the degree of applicability of the clinical trial data to patients with co-morbidities(15, 16).

The most common interventions are drug based directed at treatment of COVID-19 disease in 463 clinical trials. Though the main pathophysiology behind mortality, intubation, ICU admissions are cytokine storm and macrophage activation, nearly 50% of studies are anti-viral activity based interventions with anti-malarial accounting for 105 studies and other anti-viral drugs accounting for 76 studies(17). The main clinical rationale for use of anti-malarial agent against SARS-CoV-2 was based on invitro efficacy. Despite lack of mortality benefit and possible increased risk of adverse events with HCQ in published clinical studies till date, it is still the most commonly studied drug against SARS-CoV-2(18, 19). This calls for a need for an international repository of individual patient data and rapid assimilation of the available clinical evidence on deciding termination of potentially harmful interventions(20). Biological agents with immunosuppressive and immunomodulatory properties that have the potential to curb the cytokine storm and withspread macrophage activation are being studied with close to 100 studies ongoing [12].

The most important area for research in the current pandemic needs to be preventive studies. There are currently 72 studies with 19 vaccine based and 53 drug based preventive studies. There are currently 11 vaccines in human clinical trials. BCG vaccine is also being also studied for its proposed immunogenicity against SARS-CoV-2(21). Among the drug based preventive studies,50% are using hydroxychloroquine with various aspects being tested including daily vs weekly dosing, pre vs post exposure prophylaxis. From our review, we feel there is a greater need for more preventive strategy-based studies since most of the countries are re-opening and the proposed timeline for the pandemic to subside is 1-2 years(22). CPT use in COVID-19 disease was promoted by the potential efficacy in SARS, MERS and Ebola(23). There are currently 56 studies that are ongoing and from some of the published data it has been shown to be safe and effective. There is a need for information from larger datasets.

There are 79 studies that are assessing herbal based medicines and 42 studies that are assessing mesenchymal stem cells therapy. Majority of these studies are being done in China (%). These interventions are questionable with a potential to harm patients. There is a trend to combine these interventions with western medicines and the potential drug interactions may lead to further adverse events. One of the important aspects that this systematic review shows is the lack of enough studies on rehabilitation and mental health-related interventions. There are only 30 studies that account only for 3% of all intervention-based studies that are ongoing. Home quarantine, emotional distress from loss of loved ones, loss of job, new oxygen requirements from COVID-disease, anosmia are some of the problems faced by patients recovering from COVID-19. There is a huge need for more studies that focus on alleviating these problems. Majority of the patients with severe or critical COVID disease are in ICU. There are only 31 ongoing studies that are ICU based interventions like prone positioning, nitric oxide inhalation. There is a greater need for study of potential interventions that can improve outcomes in patients admitted in the ICU.

### Strengths of the systematic review

This is an extensive review of the ongoing clinical studies to give an insight into the various interventions being studied currently. We were also able to identify the outcomes being studied and provide inputs on the need for studies addressing definitive patient related outcomes like mortality, and need for mechanical ventilation and improvement of patients in the ICU. Our review also provides information regarding the utility of ongoing trials like HCQ based treatment, which have so far not shown benefit in larger studies, and re-assess the need for such studies and utilize resources for other interventions.

### Limitations

Newer clinicals are being rapidly initiated and enrolled into the clinical trial registries which makes it difficult for the review to be uptodate. Information regarding the status of clinical trials if they are active or have been terminated or completed is not clearly available from the databases. Though the review accounted for the most of the clinical trial registries, despite our best attempt, it may not be exhaustive enough to account for retrospective registration of all studies.

### Implications for practice

The majority of ongoing clinical trials seek to enroll patients that may not be representative of the actual population who are at risk of death and morbidity from COVID-19. There needs to be an emphasis on the rationality of the primary endpoints with need for all-cause mortality as the primary endpoint and other patient-related outcomes like need for mechanical ventilation and decreasing length of ICU stay as main secondary outcomes. There needs to be a higher rate of inclusion of patients with co-morbidities in clinical trials to reflect real world scenario for outcomes. Majority of ingoing studies are in the developed world and middle- and low-income countries are at a risk of grossly being underrepresented in the clinical trials.

## Conclusion

This systematic review identifies the spectrum of clinical trials and the therapeutic interventions that are being assessed against SARS-CoV-2. Multiple intervention based clinical studies against SARS-CoV-2 are being done throughout the world with a high concentration of clinical trials being done in the developed world. There is a high concern for redundancy of studies and under-representation of elderly and patients with comorbidities. Definite endpoints like mortality are being assessed in only one-fifth of the studies. This review provides information for researchers for rationale design of future clinical studies.

## Data Availability

All data regarding the manuscript will be available on email to the corresponding author

## Corresponding author

Bhanu Prasad Venkatesulu, Transitional year residency, Department of Internal medicine, Henry Ford hospital, Detroit, Michigan-48202, United States.

## Acknowledgements

We thank all the researchers for posting their clinical studies in the clinical trial registries.

## Footnotes

Contributors: BPV conceived the study; VT, JM and BPV designed the study; BPV and VT screened titles and abstracts for inclusion. BPV and VT extracted and analyzed data. BPV, PG, P, HP and VT formulated the preliminary draft, all authors revised for critical content. All authors approved the final manuscript. BPV guarantees for the data. The corresponding author attests that all listed authors meet authorship criteria and that no others meeting the criteria have been omitted.

## References

1. Dong E, Du H, Gardner L. An interactive web-based dashboard to track COVID-19 in real time. The Lancet Infectious diseases. 2020;20(5):533–4. Epub 2020/02/23. doi: 10.1016/s1473-3099(20)30120-1. PubMed PMID: 32087114; PubMed Central PMCID: PMCPMC7159018.

2. Wu Z, McGoogan JM. Characteristics of and Important Lessons From the Coronavirus Disease 2019 (COVID-19) Outbreak in China: Summary of a Report of 72-314 Cases From the Chinese Center for Disease Control and Prevention. Jama. 2020. Epub 2020/02/25. doi: 10.1001/jama.2020.2648. PubMed PMID: 32091533.

3. Abd El-Aziz TM, Stockand JD. Recent progress and challenges in drug development against COVID-19 coronavirus (SARS-CoV-2) - an update on the status. Infection, genetics and evolution: journal of molecular epidemiology and evolutionary genetics in infectious diseases. 2020;83:104327. Epub 2020/04/23. doi: 10.1016/j.meegid.2020.104327. PubMed PMID: 32320825; PubMed Central PMCID: PMCPMC7166307.

4. Tesia B, Vinicius A, Cleber C. M-F, Daniel K, Scott S. A, Charles S, et al. Computational Models Identify Several FDA Approved or Experimental Drugs as Putative Agents Against SARS-CoV-22020.

5. Ciliberto G, Cardone L. Boosting the arsenal against COVID-19 through computational drug repurposing. Drug discovery today. 2020. Epub 2020/04/19. doi: 10.1016/j.drudis.2020.04.005. PubMed PMID: 32304645; PubMed Central PMCID: PMCPMC7158822.

6. Rodrigo R. R. D, Dennis C. CJ, Luis P. I, Jez L. M, Douglas F. N, Timothy R. P. Repurposing FDA-Approved Drugs for COVID-19 Using a Data-Driven Approach2020.

7. Moher D, Liberati A, Tetzlaff J, Altman DG. Preferred reporting items for systematic reviews and meta-analyses: the PRISMA statement. 2009;339:b2535. doi: 10.1136/bmj.b2535 %J BMJ.

8. Treatments for COVID-19: Canadian Arm of the SOLIDARITY Trial. Available from: https://ClinicalTrials.gov/show/NCT04330690.

9. https://doi.org/10.1186/ISRCTN839711511. Public health emergency SOLIDARITY trial of treatments for COVID-19 infection in hospitalized patients. 2020.

10. Study to Evaluate the Safety and Antiviral Activity of Remdesivir (GS-5734™) in Participants With Severe Coronavirus Disease (COVID-19). Available from: https://ClinicalTrials.gov/show/NCT04292899.

11. Colchicine Coronavirus SARS-CoV2 Trial (COLCORONA). Available from: https://ClinicalTrials.gov/show/NCT04322682.

12. CROWN CORONATION: Chloroquine RepurpOsing to healthWorkers for Novel CORONAvirus mitigaTION. Available from: https://ClinicalTrials.gov/show/NCT04333732.

13. Coronavirus Response - Active Support for Hospitalised Covid-19 Patients. Available from: https://ClinicalTrials.gov/show/NCT04343001.

14. Will Hydroxychloroquine Impede or Prevent COVID-19. Available from: https://ClinicalTrials.gov/show/NCT04341441.

15. Fleming TR, Powers JH. Biomarkers and surrogate endpoints in clinical trials. Statistics in medicine. 2012;31(25):2973–84. Epub 2012/06/20. doi: 10.1002/sim.5403. PubMed PMID: 22711298; PubMed Central PMCID: PMCPMC3551627.

16. Furberg CD. To whom do the research findings apply? Heart (British Cardiac Society). 2002;87(6):570–4. Epub 2002/05/16. doi: 10.1136/heart.87.6.570. PubMed PMID: 12010948; PubMed Central PMCID: PMCPMC1767149.

17. Mehta P, McAuley DF, Brown M, Sanchez E, Tattersall RS, Manson JJ. COVID-19: consider cytokine storm syndromes and immunosuppression. Lancet (London, England). 2020;395(10229):1033–4. Epub 2020/03/21. doi: 10.1016/s0140-6736(20)30628-0. PubMed PMID: 32192578.

18. Tang W, Cao Z, Han M, Wang Z, Chen J, Sun W, et al. Hydroxychloroquine in patients with mainly mild to moderate coronavirus disease 2019: open label, randomised controlled trial. 2020;369:m1849. doi: 10.1136/bmj.m1849 %J BMJ.

19. Borba MGS, Val FFA, Sampaio VS, Alexandre MAA, Melo GC, Brito M, et al. Effect of High vs Low Doses of Chloroquine Diphosphate as Adjunctive Therapy for Patients Hospitalized With Severe Acute Respiratory Syndrome Coronavirus 2 (SARS-CoV-2) Infection: A Randomized Clinical Trial. JAMA network open. 2020;3(4):e208857. Epub 2020/04/25. doi: 10.1001/jamanetworkopen.2020.8857. PubMed PMID: 32330277.

20. Cosgriff CV, Ebner DK, Celi LA. Data sharing in the era of COVID-19. The Lancet Digital health. 2020;2(5):e224. Epub 2020/05/07. doi: 10.1016/s2589-7500(20)30082-0. PubMed PMID: 32373785; PubMed Central PMCID: PMCPMC7194831.

21. O’Neill LAJ, Netea MG. BCG-induced trained immunity: can it offer protection against COVID-19? Nature reviews Immunology. 2020:1–3. Epub 2020/05/13. doi: 10.1038/s41577-020-0337-y. PubMed PMID: 32393823; PubMed Central PMCID: PMCPMC7212510.

22. Gates B. Responding to Covid-19 - A Once-in-a-Century Pandemic? The New England journal of medicine. 2020;382(18): 1677–9. Epub 2020/02/29. doi: 10.1056/NEJMp2003762. PubMed PMID: 32109012.

23. Winkler AM, Koepsell SA. The use of convalescent plasma to treat emerging infectious diseases: focus on Ebola virus disease. Current opinion in hematology. 2015;22(6):521–6. Epub 2015/10/13. doi: 10.1097/moh.0000000000000191. PubMed PMID: 26457963.

